# Evaluation of Eye-tracking for a Decision Support Application

**DOI:** 10.1101/2020.10.19.20215087

**Authors:** Shyam Visweswaran, Andrew J King, Mohammadamin Tajgardoon, Luca Calzoni, Gilles Clermont, Harry Hochheiser, Gregory F Cooper

## Abstract

Eye-tracking is used widely to investigate attention and cognitive processes while performing tasks in electronic medical record (EMR) systems. We explored a novel application of eye tracking to collect training data for a machine learning-based clinical decision support tool that predicts which patient data are likely to be relevant for a clinical task. Specifically, we investigated in a laboratory setting the accuracy of eye tracking compared to manual annotation for inferring which patient data in the EMR are judged to be relevant by physicians. We evaluated several methods for processing gaze points that were recorded using a low-cost eye tracking device. Our results show that eye-tracking achieves accuracy and precision of 69% and 53% respectively compared to manual annotation and are promising for machine learning. The methods for processing gaze points and scripts that we developed offer a first step in developing novel uses for eye-tracking for clinical decision support.

**LAY SUMMARY:** In the context of electronic medical record systems, eye-tracking is used extensively to explore attention and cognitive processes. We investigated a novel application of eye tracking to collect training data for machine learning-based clinical decision support. We evaluated several methods for processing gaze points that were recorded using a low-cost eye tracking device. The methods for processing gaze points and scripts that we developed offer a first step in developing novel uses for eye-tracking for clinical decision support.

## 1 INTRODUCTION

With the advent of affordable eye tracking technology [1], there is growing interest in leveraging eye-tracking to support advanced types of clinical decision support (CDS) tools in electronic medical record (EMR) systems. For example, eye tracking could capture which data (e.g., vital signs, laboratory test results, medication orders, etc.) in a patient’s EMR a physician has viewed in the context of a clinical task [2]. If eye-tracking devices were deployed on EMR computer monitors then the data that are viewed by many physicians could be collected, and machine learning models derived from such data can predict which data are likely to be relevant in a given patient. Such predictive models can form the basis of a CDS tool to highlight relevant patient data and draw the physician’s attention to them [3]. Further, such a tool has the potential to mitigate the cognitive overload arising from the large amounts of patient data that physicians have to collate and assess in data-rich settings like the intensive care unit (ICU).

We developed in a Learning EMR (LEMR) system to investigate the feasibility of a CDS tool to predict and highlight relevant data [4]. The LEMR system relies on supervised machine learning models that predict which patient data are likely to be relevant in the context of a clinical task [5, 6]. However, a critical barrier to building machine learning models for a LEMR system is the acquisition of training data regarding which patient data are relevant for a clinical task. Such data are not recorded in sufficient granularity in currently deployed EMR systems. Hence, we collected training data in a laboratory setting where physicians reviewed patient cases and provided manual annotations about which data were relevant. However, manual annotation is onerous, expensive, and time-consuming, and limits the amount of training data that can be collected. Eye tracking offers an alternative method for capturing training data, and we investigated whether eye tracking data are as accurate as manual annotations. If so, eye tracking can provide a promising, higher-throughput alternative that if deployed on EMR systems will unobtrusively capture which data are viewed by thousands of physicians and provide large volumes of data for machine learning. Furthermore, the availability of inexpensive eye-tracking devices makes the broad deployment of eye-tracking enabled systems feasible.

## 2 BACKGROUND

In this section, we provide brief descriptions of eye tracking technology and its application to EMR systems, the LEMR system, and the eye-tracking device used in the LEMR system.

### 2.1 Eye tracking and its applications to EMR systems

Eye tracking is a method to track and record eye movements and gaze points across time and task, and it is commonly used for observing and measuring the allocation of visual attention [7]. Eye tracking devices record a sequence of gaze points with a regular sampling rate. Gaze point data form the basis of a variety of analyses of visual attention, such as the characterization of fixation and saccadic eye movements in an area of interest (AOI) [8] and dwell time, which is the total amount of time spent looking within an AOI [9]. Eye tracking is also widely used to study cognitive processes that underlie a variety of tasks such as visual search, comprehension, and judgment and decision-making [10].

A range of methods have been developed to measure visual attention and dwell time in AOIs from gaze point data. Simple methods calculate the dwell time or the time spent looking at an AOI by summing the time that the gaze points were within the AOI [11]. More sophisticated methods identify fixations within an AOI based on the assumption that visual attention occurs only during fixations, and calculate the dwell time by summing the time that the fixations were within the AOI [12].

In the context of EMR systems, eye tracking research has focused on understanding users and their interactions with the systems. For example, investigators have used eye-tracking to understand clinical reasoning [13], track information search patterns [14], evaluate usability [15], measure time use [16], and investigate visual and cognitive processes while performing tasks [17]. Recent reviews of the literature have described the application of eye-tracking in clinical decision-making [18] and usability of EMR systems [19]. However, little work has been done to investigate the use of eye-tracking in EMR systems to enable clinical decision support tools. In previous work, we described the application of eye-tracking to clinical decision support for the collection of training data for deriving machine learning models of relevant patient data [2, 20].

### 2.2 The Learning Electronic Medical Record (LEMR) system

We developed the LEMR system to highlight relevant patient data by using machine learning models to identify such data in the context of a clinical task for a specific patient [3, 5]. The LEMR system can also be used to collect training data for machine learning. The LEMR interface, shown in Figure 1, enables the collection of data that is judged to be relevant for a clinical task by a physician in two ways: (a) manual annotation, when the physician annotates relevant data by clicking on checkboxes (see Figure 3), and (b) eye gaze, when an eye-tracking device records gaze points while the physician is reviewing the patient’s record (see Figure 2). Our goal in developing the LEMR system was not to replicate an entire modern EMR but to have a useful prototype for displaying patient data and for capturing annotations to support studies in a laboratory setting.

**Figure 1:**
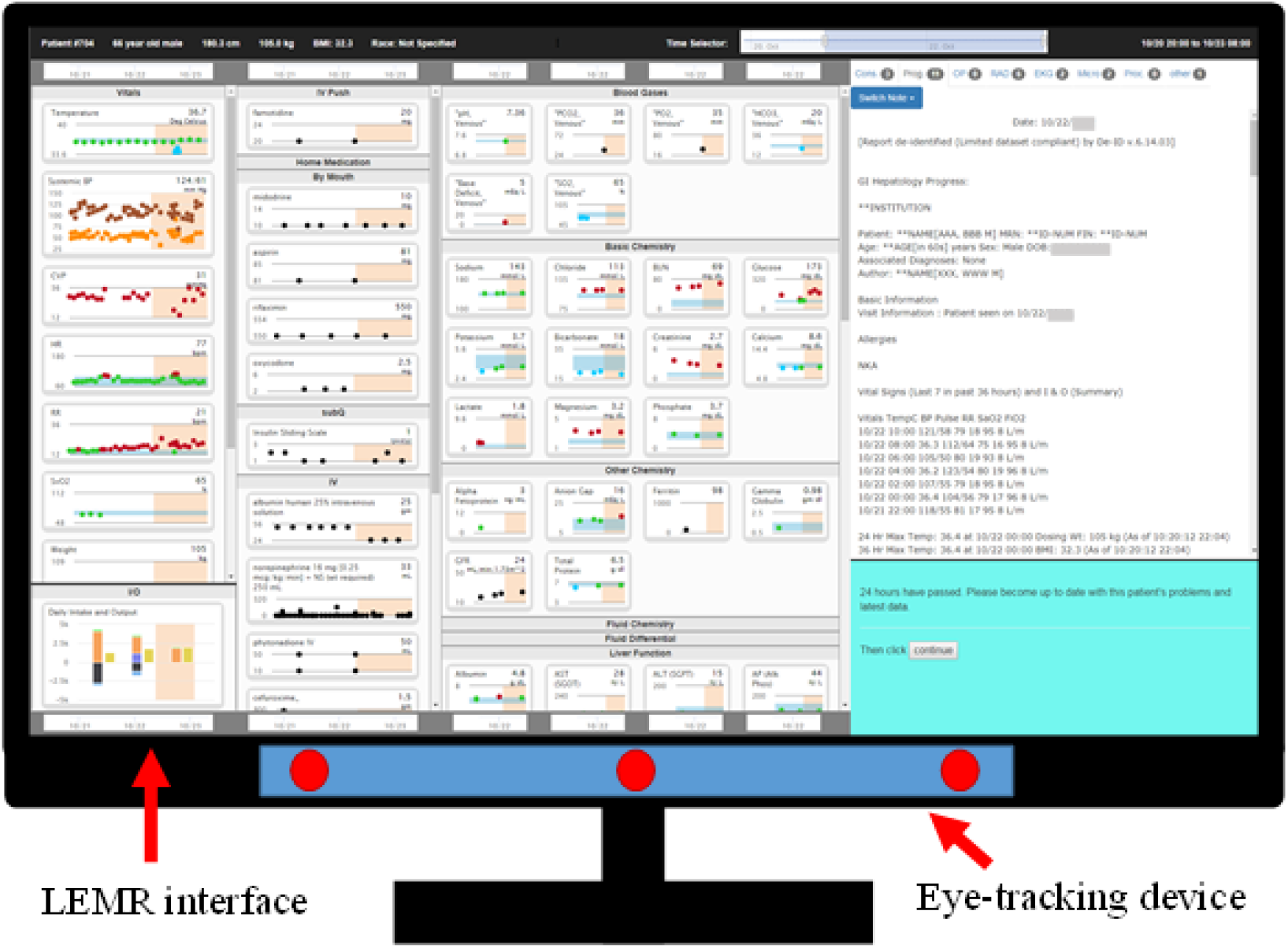
A computer monitor displaying the LEMR interface as it appears during the familiarization and preparation tasks (see Methods 3). From left to right, the system displays patient data on vital signs, ventilator settings, intake and output, medication administrations, laboratory test results, and free-text notes and reports. The eye-tracking device mounted at the bottom is used to capture gaze points during the preparation task (see Methods 3).

**Figure 2:**
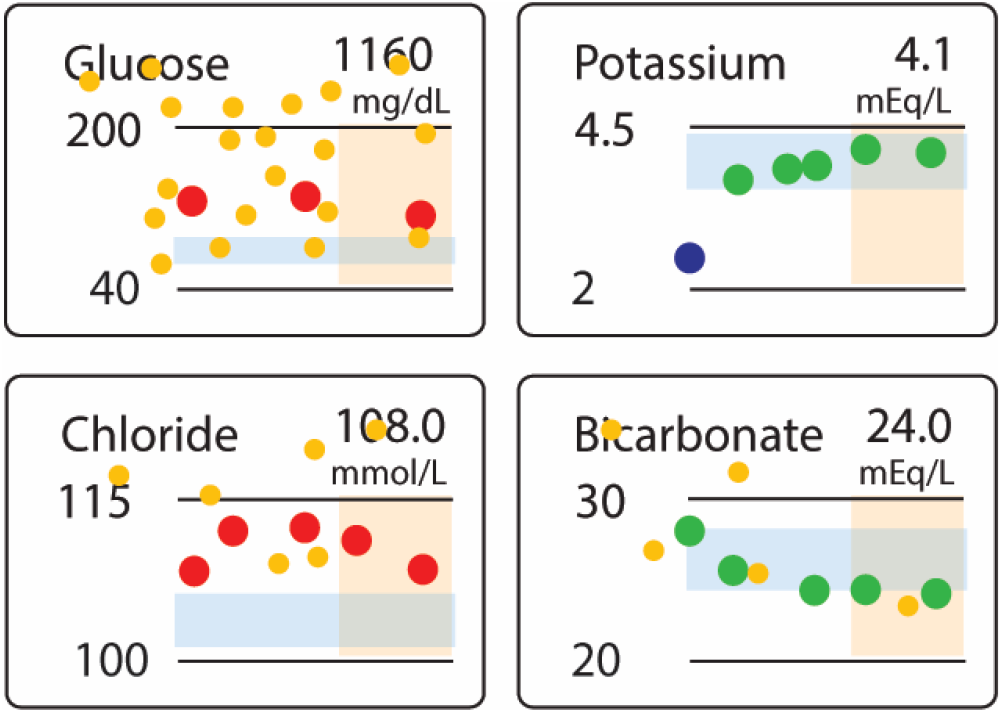
A portion of the LEMR interface as it appears during the preparation task (see Methods 3) showing four laboratory test results. The horizontal light blue band indicates the normal range for the corresponding laboratory test and the vertical light orange band indicates the most recent 24-hour period. The larger green circles, red circles, and purple circles denote normal, high, and low values of the corresponding laboratory values. The smaller orange circles denote the location of gaze points recorded by the eye-tracking device; these are shown for illustrative purposes only and are not visible on the interface.

**Figure 3:**
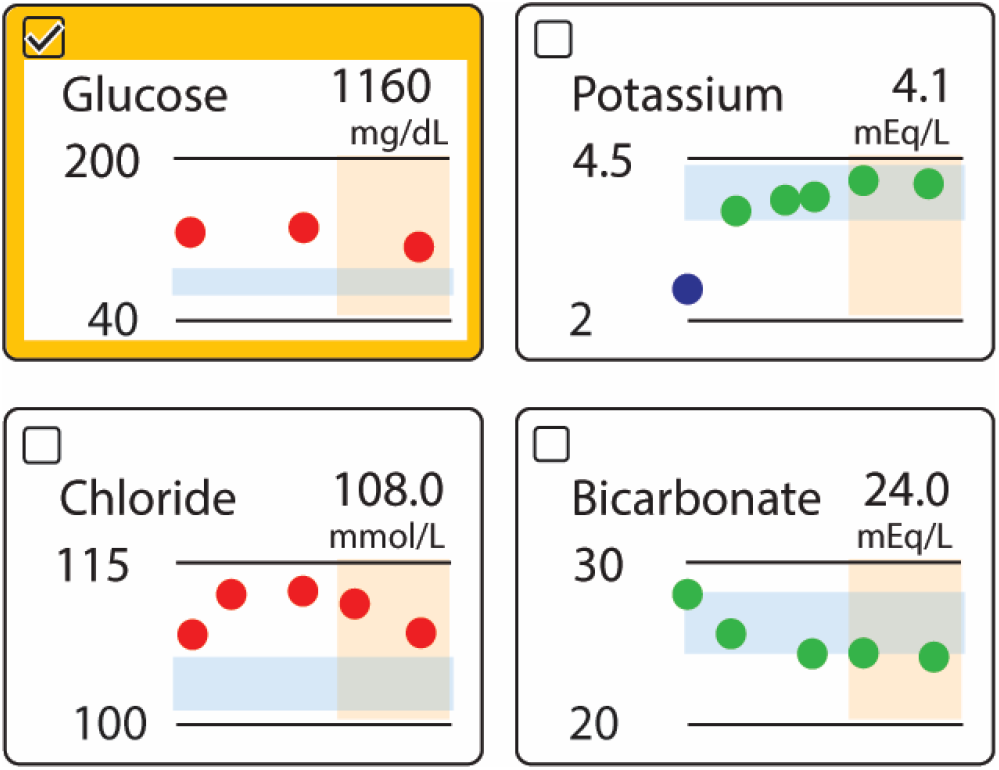
A portion of the LEMR interface as it appears during the annotation task (see Methods 3) showing four laboratory test results with checkboxes. Physicians indicate which patient data are relevant by clicking on the corresponding checkboxes. The glucose laboratory test is surrounded by a yellow margin to indicate that its checkbox has been clicked.

### 2.3 Eye-tracking device

The LEMR system is outfitted with Tobii EyeX, an inexpensive portable eye-tracking device and software package, which is primarily marketed for developing computer gaming and virtual reality applications [21]. The hardware component, the Tobii EyeX Controller, is mounted at the bottom edge of a computer monitor (see Figure 1) and samples eye gaze point coordinates at approximately 60 Hz. The software component, the Tobii EyeX Software Development Kit, records and outputs x-axis and y-axis gaze point coordinates for each eye.

## 3 METHODS

In this section we describe the study, the clinical task, data collection, the methods that we applied to process the gaze point data, and the evaluation measures we used.

### 3.1 Study description

We randomly selected 178 patients who were admitted to an ICU between June 2010 and May 2012 at the University of Pittsburgh Medical Center, and had a diagnosis of either acute kidney failure (AKF; ICD-9 584.9 or 584.5; 93 patients) or acute respiratory failure (ARF; ICD-9 518.81; 85 patients). Eleven critical care physicians reviewed the EMRs of the selected patients in the LEMR system with eye-tracking in a laboratory setting. Each patient record was reviewed by a single physician who provided both manual annotations and eye-gaze data. The number of reviewed records per physician ranged from 12 to 22 because the physicians varied in the speed of reviewing and the patient records varied in the amount of data they contained.

### 3.2 Clinical task

We chose the clinical task of identifying relevant patient data that have accumulated during the past day to present a summary of the patient’s clinical status at morning rounds in the ICU. This task is performed daily and is typically time-consuming, with the physician painstakingly searching the EMR to identify and retrieve relevant data. Each patient record was loaded into the LEMR system as shown in Figure 1. A physician reviewed a record by completing three tasks sequentially. In the familiarization task, the physician was shown patient data from the time of ICU admission up to 8:00 AM on a random ICU day between day two and the day before discharge from the ICU (inclusive). The physician was asked to review the data, become familiar with the patient and understand the clinical course. After becoming familiar with the patient, the physician switched to the preparation task, and was shown an additional 24 hours of patient data with instructions to review the new data for the task of summarizing the clinical status for presentation at morning rounds. During this task, eye-tracking was used to record the physician’s gaze points. After reviewing the new data, the physician turned to the annotation task, and indicated which patient data was relevant by clicking on checkboxes.

### 3.3 Data collection

From gaze points recorded during the preparation task, we estimated which patient data were considered to be relevant by measuring the dwell times within AOIs; from the manual checkbox annotations recorded during the annotation task, we derived a reference standard of patient data that was specified as relevant by the same physician. We compared the performance of the gaze-derived relevant patient data against the manual annotation reference standard.

### 3.4 Methods for processing gaze data

We evaluated four methods for processing gaze points to infer relevant patient data. We focused only on data on vital signs, ventilator settings, intake and output measurements, laboratory test results, and medication administration, and excluded free-text notes and reports (see Figure 1). These data are presented in rectangular areas in the LEMR interface and are the AOIs for which we applied four methods to calculate dwell times. We assume that AOIs with longer dwell times indicate relevant patient data. We selected two fixation identification algorithms that include the dispersion-threshold identification (I-DT) and area-of-interest identification (I-AOI) methods. In addition, we developed two simple gaze point algorithms called the gaze point (GP) and distributed gaze point (DGP) methods. The fixation algorithms measure the dwell times of fixations while the gaze point algorithms measure dwell times of all gaze points within a AOI. We provide brief descriptions of the four methods next.

The I-DT method identifies tightly clustered groups of gaze points as fixations. More specifically, it identifies a fixation as a collection of consecutive gaze points such that the points are within a maximum distance of one another (called dispersion) and within a period of time exceeding some minimum duration (generally 100 milliseconds) [8]. Thus, the I-DT requires two input parameters, the dispersion threshold and the duration threshold. Therefore in our experiments, we explored a range of values for the input parameters. The dispersion threshold was selected from values [50, 80, 100, 150, 200 pixels] and the duration threshold was selected from values [10, 20, 30, 40 data points]. Since the sampling frequency is 60 Hz, the interval between consecutive data points is 16.7 milliseconds; thus 10 data points for the duration threshold translates to a duration of 167 milliseconds.

The I-AOI method identifies fixations in a fashion similar to the I-DT; however, it identifies fixations that occur within one or more AOIs. I-AOI utilizes a duration threshold to distinguish fixations in the AOI from saccades crossing that area [8]. Thus, the I-AOI requires a duration threshold as an input parameter. We selected the duration threshold from values [10, 20, 30, 40, 50, 100, 150, 200 data points].

We developed two simple and computationally efficient gaze point methods that do not rely on fixation identification. The GP method maps all gaze points to AOIs without classifying the points as part of a fixation or a saccade. A higher proportion of the total recorded gaze points that map to an AOI results in a longer dwell time and indicates that more visual attention has been directed there (see Appendix A for pseudocode).

The DGP method is a probabilistic refinement of the GP method, in which each gaze point contributes to adjacent AOIs in a probabilistic fashion. The fractional contribution of a gaze point to an AOI is equal to the density of a bivariate normal distribution (see Appendix A for pseudocode). The means of the distribution are located at the center of the gaze point and the variances are derived from the average error of the eye-tracking device in the horizontal and vertical directions that we estimated in a prior study [2].

### 3.5 Performance measures

We evaluated the performance of the methods with accuracy, precision, and recall. In the context of a gaze-point processing method, a true positive is an AOI that was identified as relevant by both gaze-point processing and manual annotation, a false positive is an AOI that was identified as relevant by gaze-point processing but not by manual annotation, a true negative is an AOI that was identified as irrelevant by gaze-point processing and manual annotation, and a false negative is an AOI that was identified as irrelevant by gaze-point processing but as relevant by manual annotation. Accuracy is obtained by dividing the sum of true positives and true negatives by the total number of AOIs. Precision is obtained by dividing the number of true positives by the sum of true positives and false positives and recall is obtained by dividing the number of true positives by the sum of true positives and false negatives.

## 4 RESULTS

The reviewers were physicians trained in critical care medicine and included fellows and attending physicians. Characteristics of the reviewers are summarized in Table 1.

**Table 1:** Characteristics of physician reviewers.

| Number of physicians | Average number of years spent in ICU | Average number of weeks per year spent rounding in ICU |
| --- | --- | --- |
| 11 | 1.8 (0.3 – 7.0) | 34 (26 – 42) |

The performance of the four methods for processing gaze data is shown in Table 2. Overall, GP had the highest accuracy and precision of 69% and 53% respectively and DGP had the highest recall at 48%. I-DT and I-AOI both had lower values on precision and recall and slightly lower accuracy than GP and DGP.

**Table 2:** Accuracy, precision, and recall values with standard error of four methods for processing gaze data. The highest values for each performance measure are in bold font.

| Method | Accuracy (%) | Precision (%) | Recall (%) |
| --- | --- | --- | --- |
| **I-DT** | 67 ± 0.03 | 46 ± 0.05 | 26 ± 0.23 |
| **I-AOI** | 68 ± 0.03 | 49 ± 0.09 | 31 ± 0.26 |
| **GP** | **69 ± 0.04** | **53 ± 0.10** | 38 ± 0.28 |
| **DGP** | 67 ± 0.05 | 50 ± 0.08 | **48 ± 0.25** |

## 5 DISCUSSION

We evaluated four methods for processing gaze data obtained to infer what patient data physicians judged to be relevant for summarizing the patient’s clinical status at morning rounds in the ICU. Compared to manual annotation, the results support the use of eye tracking and relatively simple methods for processing gaze data to infer data relevance with modest accuracy and precision. We derived machine learning models using gaze data to predict relevant patient data in the LEMR system and found that they performed as well as models that were derived using manual annotation. The detailed results are reported in a separate publication [20]. The scripts for the four eye-tracking methods with accompanying documentation are freely available on GitHub at https://github.com/ajk77/EyeBrowserPy.

The eye tracking device, Tobii EyeX, was developed as an inexpensive device for gaming applications. It is simple to install on a computer monitor, unobtrusive, and easy to calibrate. The Software Development Kit provides API bindings for several programming languages including Python that are straightforward to use for programming.

There are several limitations to our study. A key limitation is that the methods we used infer visual attention or seeing rather than cognition, and seeing does not imply that the information was cognitively processed. A second limitation is that the eye tracking device, Tobii EyeX, was not developed explicitly for research applications and the device has modest temporal and spatial resolution and sampling frequency. However, it was adequate for our application that only required monitoring of simple eye movements. Further, the device could not track head movements and the reviewers in our study had to restrain their head movements. However, this limitation may be mitigated with newer devices such as Tobii Eye Tracker 5 that are capable of tracking both head and eye movements and offer the ability to robustly estimate the coordinates of eye-gaze even if the head position changes [22]. A third limitation is that the interface of the LEMR system is significantly different from the vendor EMR systems currently used in clinical care, and furthermore may not be optimal for the review of patient data. Further studies are needed to assess and improve the LEMR interface. A fourth limitation is that given our results of modest accuracy and precision of eye-tracking, the performance of the models derived from such data may be imperfect and unreliable to such an extent that it leads to poorer performance and trust in the LEMR system [23, 24]. One approach to mitigating this limitation that we plan to investigate in future studies is to examine whether the performance of the models can be improved with a combination of smaller amounts of more accurate, manually obtained data with larger amounts of less accurate eye-tracking data over either type of data alone.

## 6 CONCLUSION

Eye-tracking provides an automated and unobtrusive method to capture which patient data physicians judge to be relevant for a specific clinical task. Gaze point data recorded with an inexpensive eye-tracking device have modest accuracy and precision in inferring relevant data and are promising for deriving machine learning models that identify and highlight relevant patient data. The methods for processing gaze points and scripts that we developed offer a first step in developing novel uses for eye-tracking for clinical decision support.

## Data Availability

The data underlying this article cannot be shared publicly due to the privacy of individuals that participated in the study as well as the privacy of the patients whose data were used in the study. The data will be shared on reasonable request to the corresponding author.

## CONTRIBUTORSHIP STATEMENT

S.V. conceived and designed the study, participated in data analysis and interpretation, drafted the manuscript and approved the final version for submission. A.J.K. conceived and designed the study, participated in data collection, analysis and interpretation, drafted the manuscript and approved the final version for submission. M.T. participated in data analysis and interpretation, drafted the manuscript and approved the final version for submission. L.C. made critical revisions to the manuscript and approved the final version for submission. G.C. participated in data collection, made critical revisions to the manuscript and approved the final version for submission. H.H. participated in data analysis and interpretation, made critical revisions to the manuscript and approved the final version for submission. G.F.C. conceived and designed the study, participated in data analysis and interpretation, made critical revisions to the manuscript and approved the final version for submission.

## ACKNOWLEDGMENTS

This research was supported in part by the University of Pittsburgh Center for Research Computing (CRC) through the resources provided. The authors thank Ms. Yu Zhang for assistance with data preprocessing. The study was approved by the University of Pittsburgh IRB under protocol PRO14020588.

## A Appendix

**Figure A.1:**
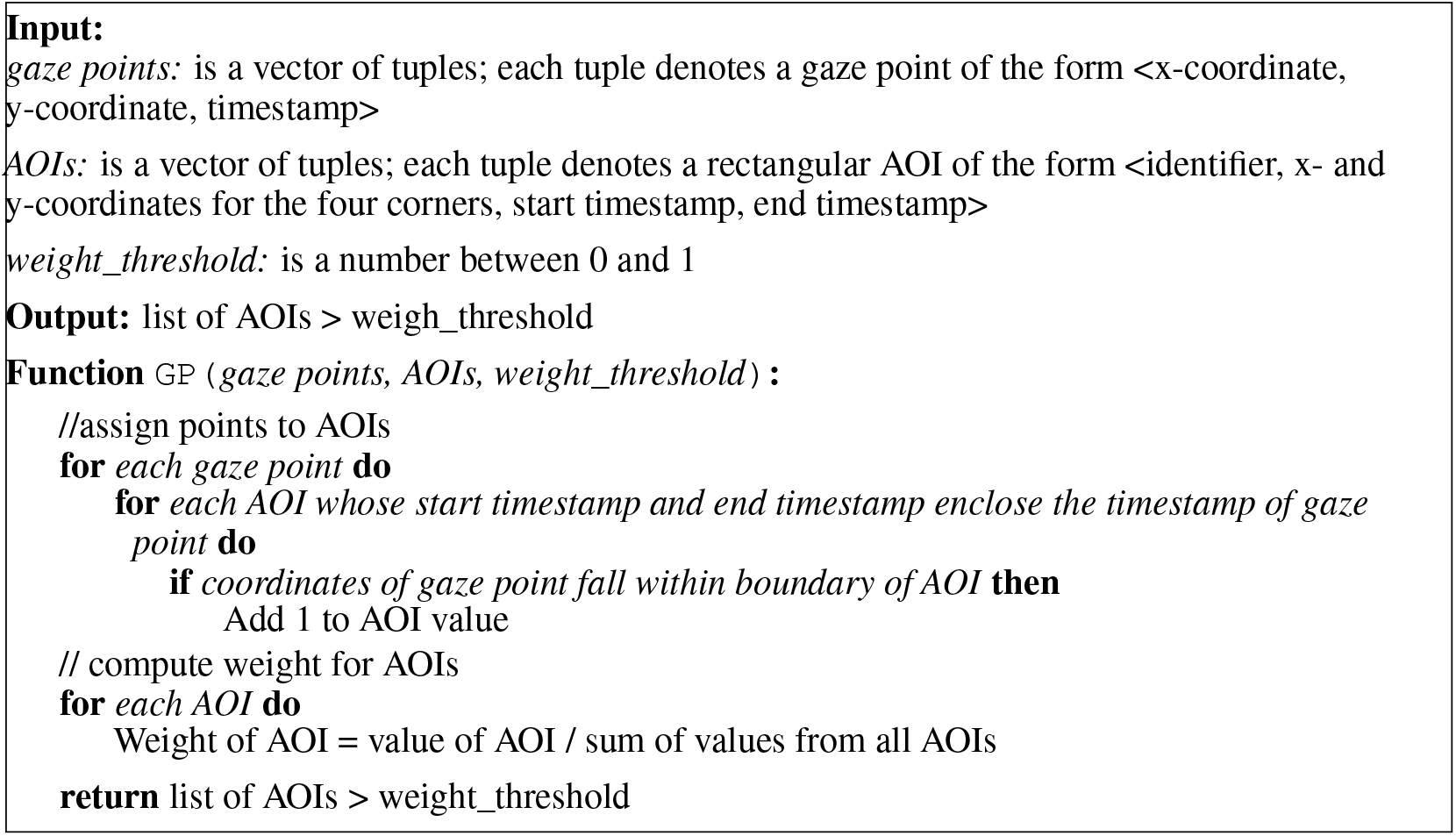
Pseudocode for the gaze point (GP) method. AOI refers to area of interest.

**Figure A.2:**
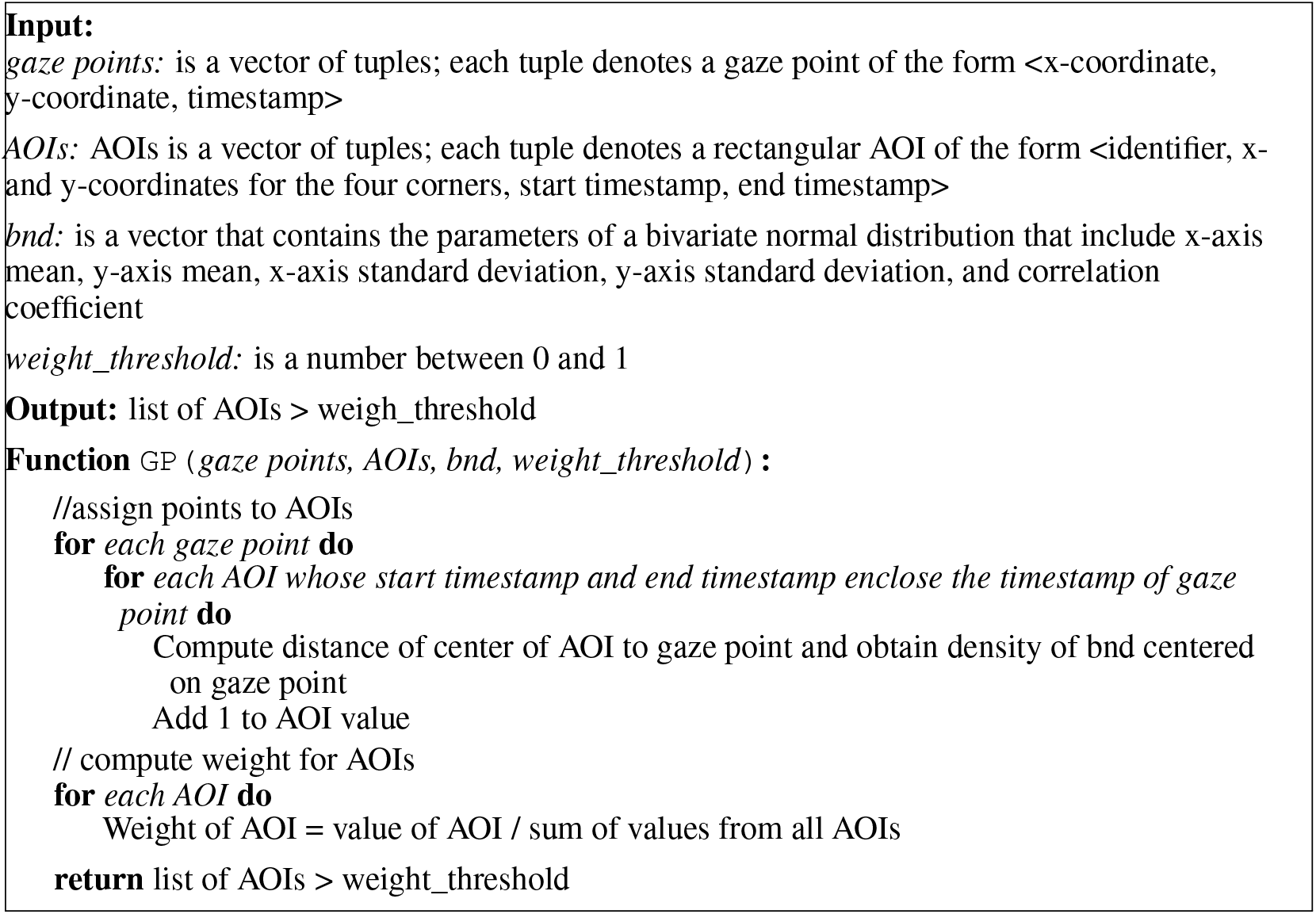
Pseudocode for the distributed gaze point (DGP) method. AOI refers to area of interest.

